# Hypomethylation of contracted D4Z4 repeats in facioscapulohumeral muscular dystrophy

**DOI:** 10.1101/2022.06.24.22276750

**Authors:** Yosuke Hiramuki, Yuriko Kure, Yoshihiko Saito, Megumu Ogawa, Keiko Ishikawa, Madoka Mori-Yoshimura, Yasushi Oya, Yuji Takahashi, Dae-Seong Kim, Noriko Arai, Chiaki Mori, Tsuyoshi Matsumura, Tadanori Hamano, Kenichiro Nakamura, Koji Ikezoe, Shinichiro Hayashi, Yuichi Goto, Satoru Noguchi, Ichizo Nishino

**Affiliations:** Department of Neuromuscular Research, National Institute of Neuroscience, National Center of Neurology and Psychiatry, Kodaira, Japan; Medical Genome Center, National Center of Neurology and Psychiatry, Kodaira, Japan; Department of Neurology, National Center Hospital, National Center of Neurology and Psychiatry, Kodaira, Japan; Department of Neurology, Pusan National University Yangsan Hospital, Yangsan, Republic of Korea; Department of Neurology and Cerebrovascular Medicine, Saitama Medical University International Medical Center, Saitama, Japan; Department of Neurology, National Hospital Organization Osaka Toneyama Medical Center, Osaka, Japan; Second Department of Internal Medicine, Division of Neurology, Department of Aging and Dementia, Faculty of Medical Sciences, University of Fukui, Fukui, Japan; Department of Neurology, National Hospital Organization Nishi-Beppu National Hospital, Beppu, Japan; Department of Neurology, Matsuyama Red Cross Hospital, Matsuyama, Japan; Department of Mental Retardation and Birth Defect Research, National Institute of Neuroscience, National Center of Neurology and Psychiatry, Kodaira, Japan

**Keywords:** CpG methylation, CRISPR/Cas9, Facioscapulohumeral muscular dystrophy, D4Z4, DUX4, Nanopore sequencer

## Abstract

Facioscapulohumeral muscular dystrophy (FSHD) can be subdivided into two types: FSHD1, caused by contraction of the D4Z4 repeat on chromosome 4q35, and FSHD2, caused by mild contraction of the D4Z4 repeat plus aberrant hypomethylation mediated by genetic variants in *SMCHD1, DNMT3B*, or *LRIF1*. Genetic diagnosis of FSHD is challenging because of the complex procedures required. Here, we applied Nanopore CRISPR/Cas9-targeted resequencing for the diagnosis of FSHD by simultaneous detection of D4Z4 repeat length and methylation status at nucleotide level in genetically-confirmed and suspected patients. We found significant hypomethylation of contracted D4Z4 repeats in FSHD1 and strong correlation between methylation rate and patient phenotype. This finding can explain how repeat contraction contributes to disease pathogenesis by activating DUX4 expression.

## Introduction

Facioscapulohumeral muscular dystrophy (FSHD) is an autosomal disease characterized by muscle weakness that initially manifests in the face, shoulder, and upper arms, followed by asymmetric involvement of other muscles (Greco et al., 2020). *DUX4* is a causative gene for FSHD and is located within an approximately 3.3 kb repeat sequence, referred to as D4Z4, which comprises 1–100 repeat units (RUs) on the subtelomeric regions of chromosomes 4 and 10. Chromosome 4 has two haplotypes distal of the D4Z4 repeat, 4qA and 4qB, where only the 4qA allele contributes to FSHD development, due to the presence of a polyadenylation signal in the most distal D4Z4 RU (Lemmers et al., 2002, 2010).

FSHD has two types, FSHD1 and FSHD2, both caused by genetic defects leading to aberrant DUX4 expression in skeletal muscle (Snider et al., 2010). FSHD1 is mediated by contraction of the D4Z4 4qA allele to 1–10 RUs (Wijmenga et al., 1992), while FSHD2 is caused by a combination of milder D4Z4 contraction (8–20 RUs) and genetic variants in *SMCHD1, DNMT3B*, or *LRIF1*, which each encode epigenetic modifiers (Van Den Boogaard et al., 2016; Hamanaka et al., 2020; Lemmers et al., 2012). DNA methylation and histone modification at D4Z4 RUs are altered in FSHD (Haynes et al., 2018; Van Overveld et al., 2003; Zeng et al., 2009). CpG methylation is specifically decreased at the contracted D4Z4 repeat on chromosome 4 in FSHD1, while the D4Z4 repeats on both chromosomes 4 and 10 are hypomethylated in FSHD2 (de Greef et al., 2009; Jones et al., 2014; Van Overveld et al., 2003); however, the distribution of methylation throughout the full D4Z4 repeat sequence has not been analyzed.

Southern blotting, bisulfite sequencing, molecular combing, and next-generation sequencing are currently used for genetic diagnosis of FSHD (Zampatti et al., 2019), but these diagnostic procedures and interpretation of their results present several difficulties. First, interpretation of hybridization patterns generated by Southern blotting is complicated by the fact that the detecting probe also recognizes an additional locus on chromosome 10q that is almost completely homologous to the target 4q35 locus.

Second, two subtelomeric variations distal to D4Z4 have been identified on chromosome 4, referred to as the 4qA and 4qB alleles, and selective identification of contracted 4qA repeats is necessary, as only 4qA is associated with FSHD. Third, analysis of CpG methylation by bisulfite sequencing has been performed across the entire D4Z4 units at both the 4q and 10q loci; however, a focal region of extreme demethylation has been reported (Hartweck et al., 2013). Additionally, several patients with milder D4Z4 contraction and CpG hypomethylation have been identified, making diagnosis difficult.

Here, we applied Nanopore CRISPR/Cas9-targeted resequencing (nCATS) to measure the number of D4Z4 RUs and their methylation status in patients with FSHD. We specifically analyzed D4Z4 RUs derived from 4qA and measured the CpG methylation rate in each RU. D4Z4 RUs from 10q were also analyzed.

## Results

### Determination of numbers of D4Z4 RUs in patients with facioscapulohumeral muscular dystrophy by Nanopore sequencing

For CAS9 cleavage, we designed two types of guide RNA each for the p13E-11 (CR1/CR2) and A-haplotype (CR3/CR4) regions; the distal guides, CR3 and CR4, specifically recognized the 4qA and 10q loci, but not 4qB (Figure 1A and 1B). To validate the nCATS assay, we analyzed five samples (Sample 1–5) from patients genetically diagnosed with FSHD1 by Southern blotting (Table 1 and 2). Reads derived from the 4qA locus were obtained after alignment to the reference sequence, and the number of D4Z4 RUs were calculated from the read length (Figure 1C; red dots, Figure 1D; Supplemental Table 1). Sample 1, 2, 3, 4, and 5 carried 1, 2, 3, 4, and 5 D4Z4 RUs, respectively, consistent with results from Southern blotting.

**Figure 1.**
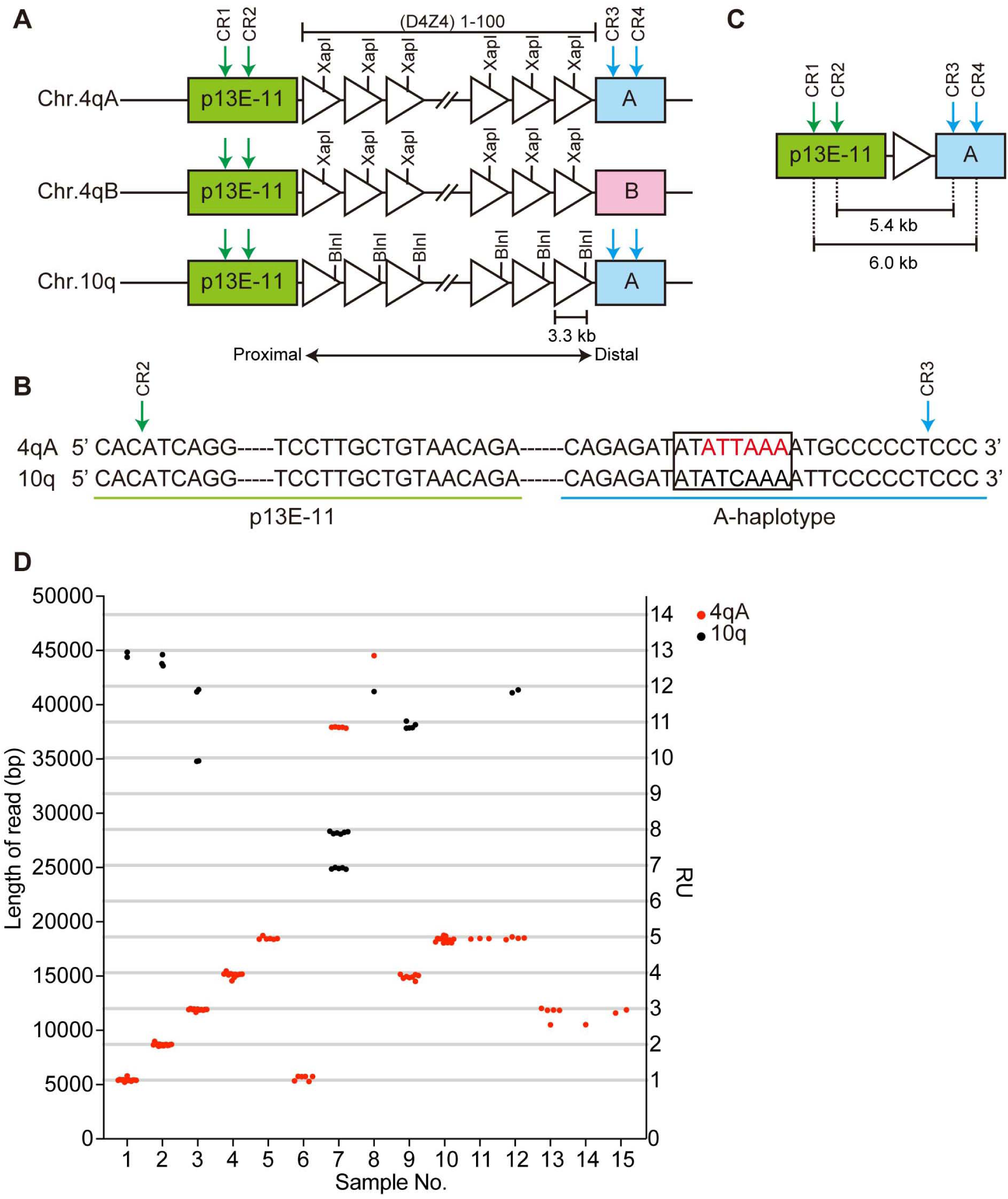
Determination of numbers of D4Z4 RUs in patients with facioscapulohumeral muscular dystrophy by Nanopore sequencing. (A) Schematic showing the D4Z4 repeat regions at the human chromosome 4qA, 4qB, and 10q loci. D4Z4 RUs are represented by triangles. The XapI and BlnI restriction enzyme sites are unique to chromosomes 4 and 10, respectively. The p13E-11, A-type haplotype, and B-type haplotype regions are indicated in green, blue, and pink, respectively. Green and blue arrows indicate crRNA cleavage sites (CR1/CR2/CR3/CR4). (B) The CR2 site in p13E-11 and CR3 site in the A-haplotype on the 4qA and 10q allele are shown by green and blue arrows, respectively. The 4qA polyadenylation signal is indicated in red. Sequences in the rectangle were used to distinguish reads from the 4qA and 10q loci. (C) Fragments carrying a single D4Z4 RU produced by CR2/CR3 or CR1/CR4 cleavage were 5.4 and 6.0 kb, respectively. (D) The length of identified reads and numbers of D4Z4 RUs are plotted. Red and black dots indicate reads derived from the 4qA and 10q loci, respectively.

**Table 1.** Patient clinical information

| Sample ID | Patient ID | Genetic diagnosis | Age ranges at hospital inspection (years) | Sex | Age ranges at onset (years) | Asymmetric weakness | Facial weakness | Scapula weakness | Humeral weakness | Beevor's sign | Other symptoms | Serum CK (IU/L) |
| --- | --- | --- | --- | --- | --- | --- | --- | --- | --- | --- | --- | --- |
| 1, 6 | 1 | FSHD1 | 11-15 | M | Birth | + | + | + | + | No data | Severe hearing loss | 935 |
| 2 | 2 | FSHD1 | 56-60 | M | 11-15 | + | + | + | + | - | - | 87 |
| 3 | 3 | FSHD1 | 11-15 | M | 11-15 | + | + | + | + | + | - | 786 |
| 4 | 4 | FSHD1 | 15-20 | M | 10-15 | + | + | + | + | + | - | 986 |
| 5 | 5 | FSHD1 | 51-55 | M | 16-20 | + | + | + | + | - | - | 288 |
| 7 | 6 | Suspected FSHD2 | 26-30 | M | 16-20 | + | + | + | + | No data | - | 1195 |
| 8 | 7 | Suspected FSHD2 | 41-45 | F | 41-45 | + | + | + | + | - | Mild hearing loss | 380 |
| 9 | 8 | Suspected FSHD1 | 16-20 | M | 11-15 | + | + | + | + | - | - | 887 |
| 10 | 9 | Suspected FSHD1 | 61-65 | F | 41-45 | + | + | + | + | + | - | 259 |
| 11 | 10 | Suspected FSHD1 | 71-75 | F | 46-50 | + | + | + | + | - | Mild hearing loss | 156 |
| 12 | 11 | Suspected FSHD1 | 11-15 | M | 11-15 | + | - | + | + | - | - | 1262 |
| 13 | 12 | Suspected FSHD1 | 21-25 | F | Childhood | + | + | + | + | No data | - | 241 |
| 14 | 13 | Suspected FSHD1 | 16-20 | F | Childhood | + | + | + | + | + | - | 267 |
| 15 | 14 | Suspected FSHD1 | 66-70 | F | 11-15 | + | + | + | + | + | - | 462 |

**Table 2.**
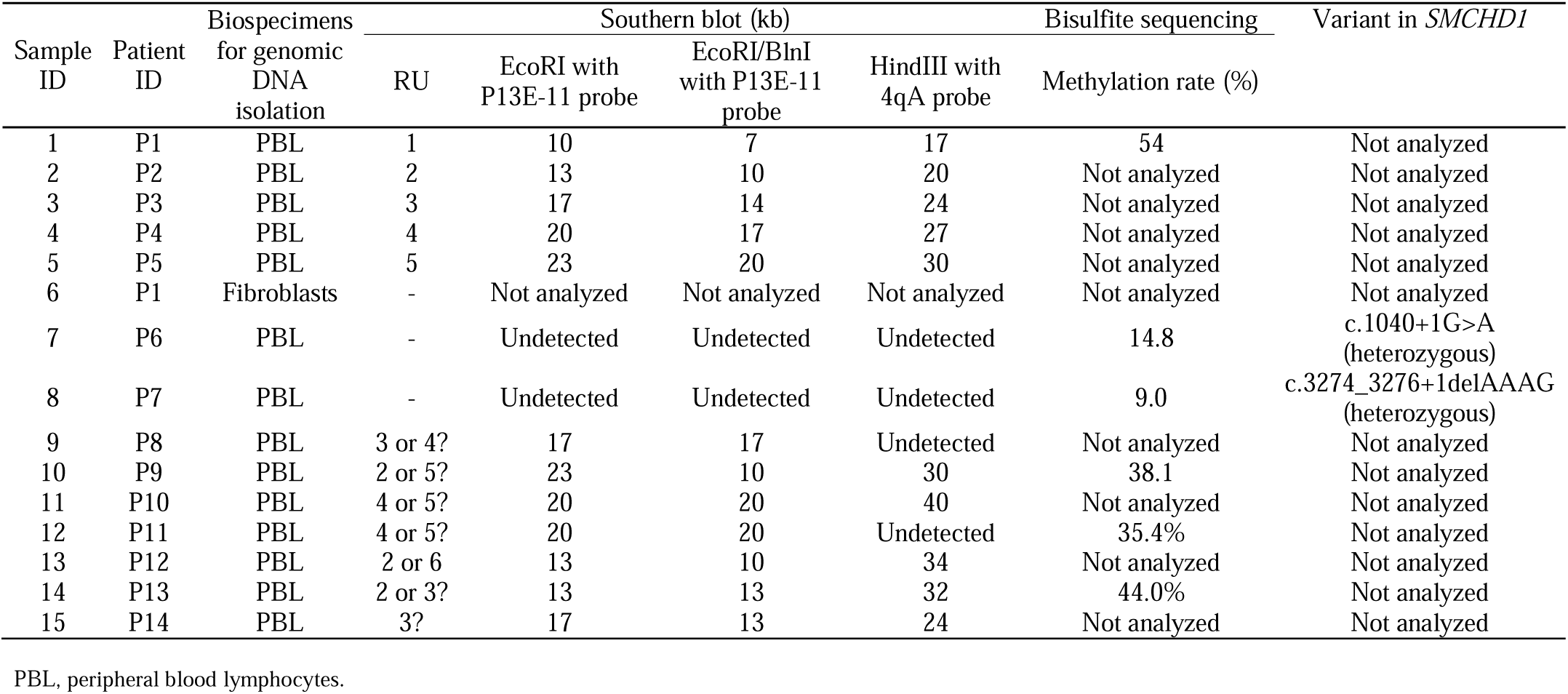
Biospecimens and results of routine genetic analyses

In addition to the 4qA locus, we also occasionally obtained reads from chromosome 10q in Samples 1 (13 RUs), 2 (13 RUs), and 3 (10 RUs and 12 RUs) (black dots in Figure 1D and Supplemental Table 2). We confirmed that both the 4qA- and 10q-derived reads were correctly assigned by identifying 4qA-specific (XapI, Non-BlnI, and pA) and 10q-specific (Non-XapI, BlnI, and Non-pA) sequences, along with the common p13E-11 sequence (Supplemental figure 1). Moreover, we confirmed that identical results were obtained using genomic DNA samples from the same subject from different sources by comparing Samples 1 and 6. These results suggest that our method enables precise determination of the number of D4Z4 RUs and the haplotypes on which the repeats reside.

We also analyzed samples that were undiagnosed by Southern blotting following linear gel electrophoresis because we failed to detect 4qA-derived bands (Samples 7 and 8) or failed to determine repeat lengths based on restriction fragment sizes (Samples 9–15) (Table 2). Using nCATS, we successfully determined the repeat lengths of 4qA-derived reads even from these challenging samples, as follows: Sample 7, 11 RUs; Sample 8, 13 RUs; Sample 9, 4 RUs; Sample 10, 5 RUs; Sample 11, 5 RUs; Sample 12, 5 RUs; Sample 13, 3 RUs; Sample 14, 3 RUs; and Sample 15, 3 RUs (Figure 1D).

### A genomic deletion detected in patients with contracted D4Z4 repeats

Interestingly, we also detected a genomic deletion, as an atypical cause of rearrangement of D4Z4 repeats. Samples 13 and 14 each generated one read with an intermediate size between 2 and 3 RUs. Sequence analysis revealed that both reads contained a deletion spanning 1.3 kb from 469 bases proximal to the most proximal D4Z4 RU to 859 bases within it (Figure 2). Deletion within D4Z4 repeats has not been reported previously in FSHD1.

**Figure 2.**
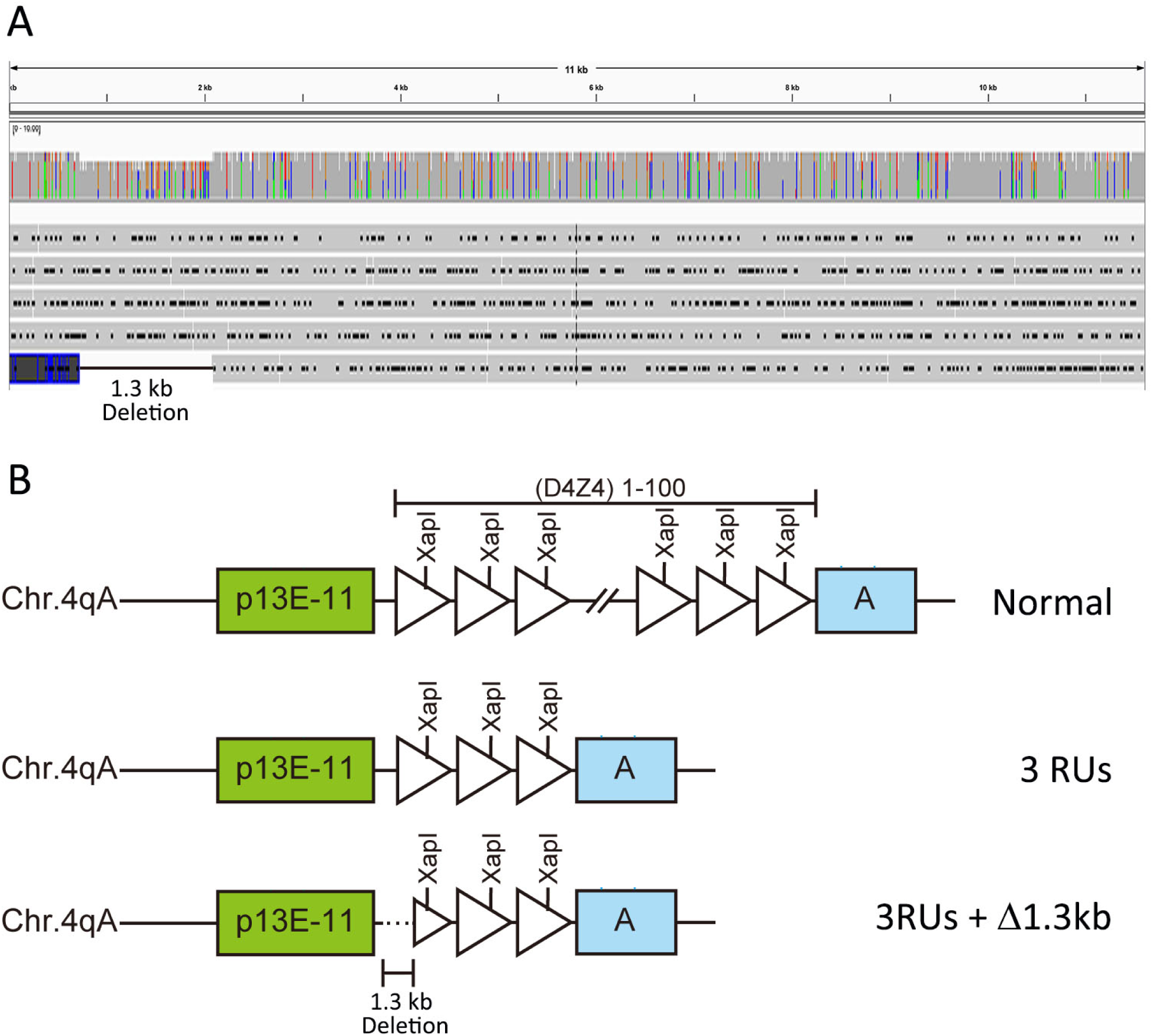
A genomic deletion detected in patients with contracted D4Z4 repeats. (A) Representative data showing reads obtained from Sample 13 mapped using Integrative Genomics Viewer. Five reads with three repeat units (RUs) (11 kb), four reads with no deletion, and one read with a 1.3 kb deletion were obtained. (B) The deletion was localized downstream of p13E-11 and extended to the middle of the most proximal D4Z4 RU.

### CpG methylation rates in D4Z4 RUs

We also used nCATS results to determine the CpG methylation status of individual reads; therefore, we calculated CpG methylation rates for each RU in 4qA and 10q-derived reads (Figure 3 and Supplemental Table 3). In FSHD1, the methylation rates of contracted 4qA-reads were consistently low, although those of the most distal D4Z4 RU at position 1 were relatively higher in most reads (Figure 3). By contrast, the methylation rates of 10q-derived reads were low in proximal RUs, but elevated toward distal RUs. Further, in FSHD2, the CpG methylation rates of both 4qA- and 10q-reads were low throughout, with the exception of a few reads, in which the most distal RU1 was relatively highly methylated.

**Figure 3.**
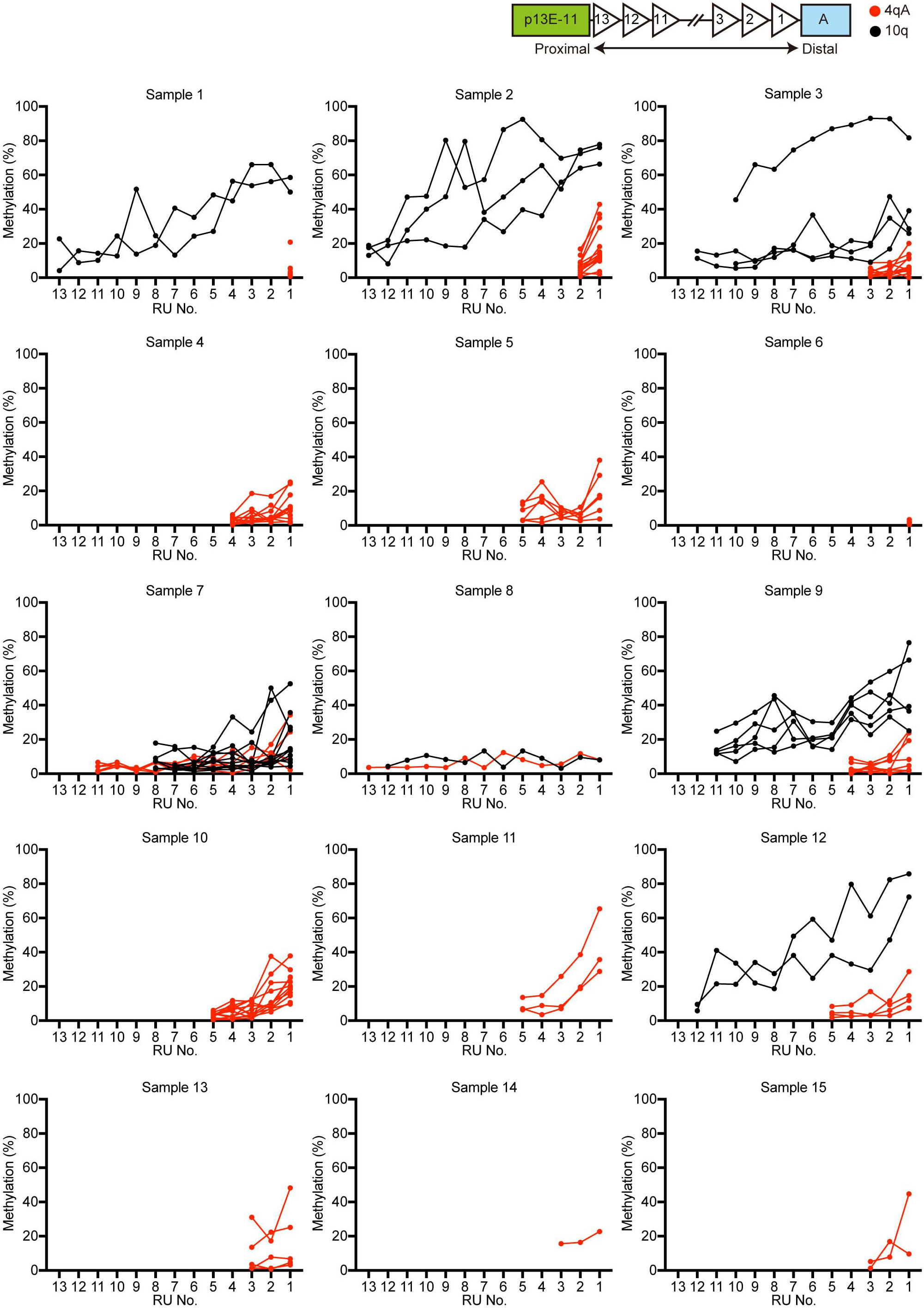
CpG methylation rates in D4Z4 Rus. CpG methylation rates of D4Z4 RUs in individual reads from the 4qA and 10q loci are plotted in red and black, respectively. D4Z4 RUs are numbered from the distal D4Z4 region.

### Methylation rates in the promoter region and gene body of the most distal D4Z4 RU

Next, we analyzed the CpG methylation rates of the promoter region and gene body of the most distal D4Z4 RU (RU1) separately (Figure 4). Although Samples 1 and 6, which contained only one RU, showed similar CpG methylation rates in the promoter region and gene body, the methylation rates of promoter regions were generally lower than those in the gene body in all other samples from patients with both FSHD1 and FSHD2.

**Figure 4.**
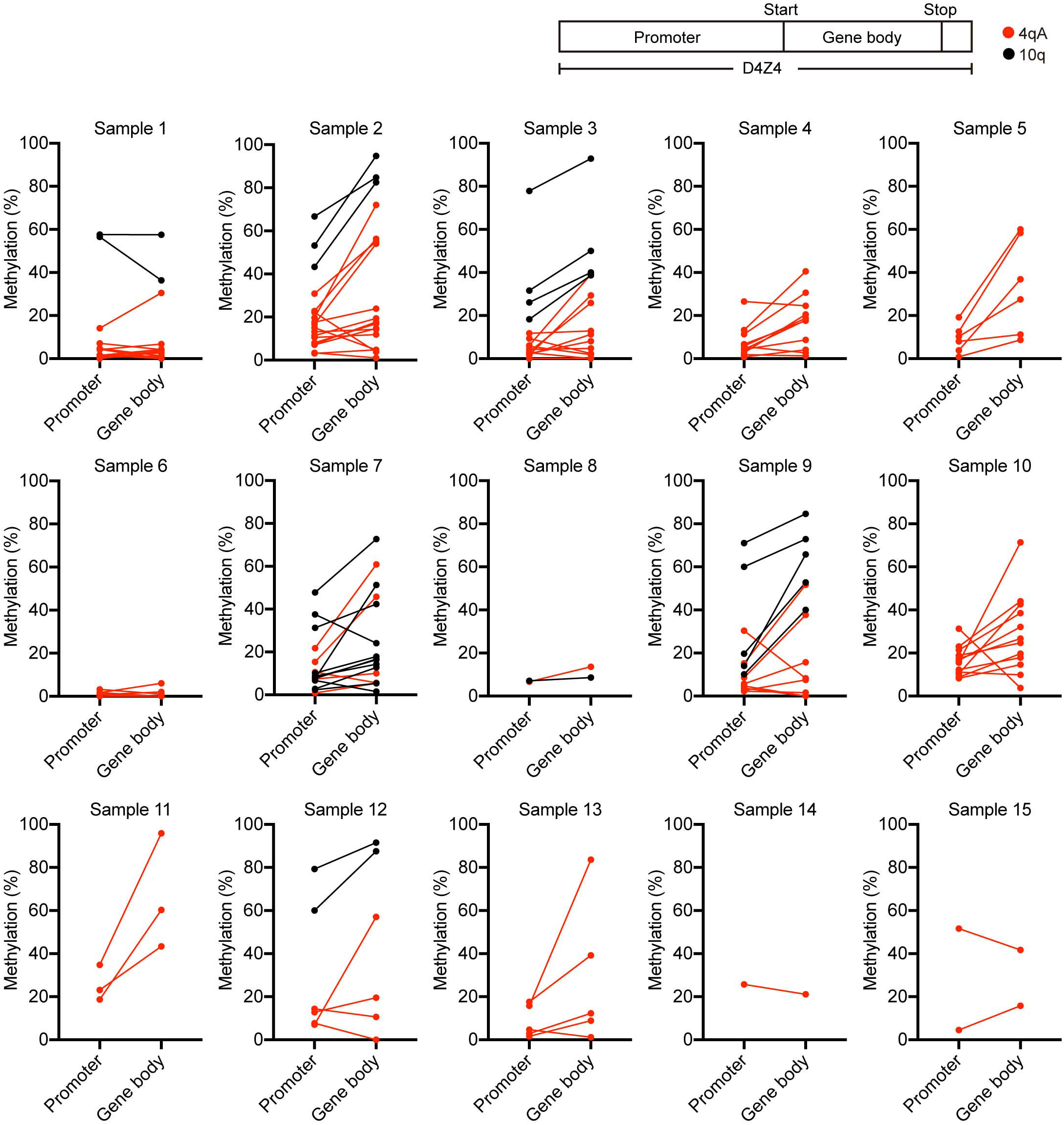
Methylation rates in the promoter region and gene body of the most distal D4Z4 RU. CpG methylation rates in the promoter and gene body of the most distal D4Z4 RU in individual reads are plotted. Reads from 4qA and 10q are shown in red and black, respectively.

### Correlation between CpG methylation rate in distal D4Z4 and patient phenotypes

Epigenetic changes in the contracted D4Z4 repeats on chromosome 4qA have been observed previously and are considered to be associated with the development of FSHD1 (de Greef et al., 2009; Jones et al., 2014; Van Overveld et al., 2003). We hypothesized that the CpG methylation rate of the most distal D4Z4 RUs is a determinant of disease development; therefore, we examined the correlation between average methylation rate of the most distal three RUs (Figure 3) and patient age at onset or at hospital inspection. As shown in Figure 5, we found a strong correlation between CpG methylation and age at onset (R^2^ = 0.645) than that between D4Z4 repeat length and age at onset (R^2^ = 0.401). Although the correlation coefficient between CpG methylation and age at hospital inspection was not high (R^2^ = 0.306), there was a tendency toward correlation, in that CpG methylation rate < 10% was associated with hospital inspection at a younger age (_≤_ 20 years old), while CpG methylation rates of 10–20% were associated with that at > 40 years old.

**Figure 5.**
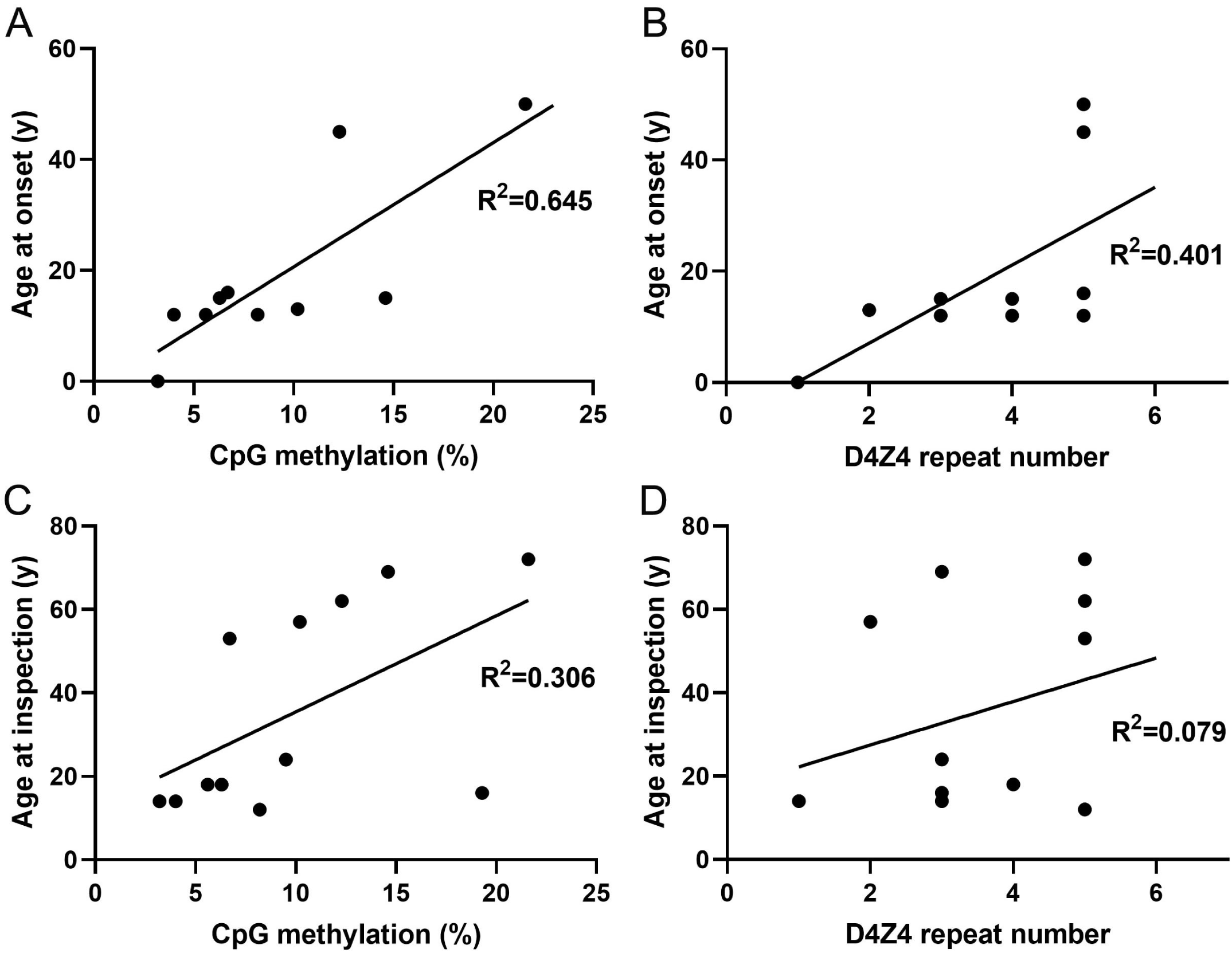
Correlation between CpG methylation rate in distal D4Z4 or repeat length, and patient phenotypes. (A and C) Scatter plots of mean CpG methylation rate in distal D4Z4 repeat units and age at disease onset (A) or age at hospital inspection (C). (B and D) Scatter plots of D4Z4 repeat number and age at disease onset (B) or age at hospital inspection (D). Correlation coefficients were calculated by linear regression

## Discussion

Nanopore sequencing was previously applied for analysis of FSHD using a bacterial artificial chromosome clone containing 13 D4Z4 repeat units (Mitsuhashi et al., 2017). In this study, we developed a direct sequencing system using nCATS to analyze clinical samples from patients with FSHD. Our system has several advantages. First, long read sequencing can be applied to analysis of a similar DNA fragment size range to that detected by Southern blotting. Second, CRISPR/CAS9 enrichment allows barcoding sequencing of five samples simultaneously, saving time and cost. Third, single-molecule sequencing technology provides genetic information at the base level and can determine the number of RUs, even in samples that have mutated restriction enzyme sites, which prevent determination of RU number by the standard Southern blotting method. Finally, the nCATS system allows simultaneous detection of CpG methylation and D4Z4 RUs numbers, providing information about local epigenetic modification of D4Z4 repeats, due to the application of single-molecule sequencing of unamplified genomic DNA molecules derived from individual nuclei, without any bias. Nevertheless, the nCATS method also has limitations. The number of sequencing reads containing mildly contracted D4Z4 repeats (11–13 RUs) detected was quite low, particularly as only a few reads were obtained from the normal 10q locus, and no reads were obtained from some samples. The reasons why we could not obtain read from chromosome 10 in all samples are; 1) the difficulty to obtain longer DNA fragments beyond 13 RUs, because we used only the reads harboring full-length D4Z4 repeat in our analysis, 2) the efficacy of CAS9 cleavage of hypermethylated DNA, because distal D4Z4 were extremely higher methylation rates. Technical improvements are required to overcome this shortcoming.

Along with successful determination of D4Z4 RU numbers in patients, we also detected atypical rearrangement of D4Z4 repeats. As shown in Figures 1D and 2, two reads of intermediate size had a 1.3 kb deletion in the most proximal D4Z4 RU, while p13E-11 was not deleted. This deletion is unlikely to be associated with the contraction of D4Z4 repeats in FSHD1, as the pathogenic alleles in FSHD1 usually maintain the intact RU structure, even when they contracted. Common atypical rearrangements found in individuals with FSHD1 have been reported, including D4Z4 proximally extended deletion (DPED1–7) alleles, which span 5.9–45.7 kb proximal to and within D4Z4, including p13E-11. In some DPED alleles, genetic elements, such as *DUX4C, FRG2, DBE-T*, and myogenic enhancers, are deleted, suggesting that their role in FSHD pathogenesis requires reevaluation (Lemmers et al., 2022).

Although the limitations of our study include the relatively small number of reads obtained from D4Z4 repeats with more than 10RUs, and the lack of analysis of the D4Z4 region derived from the 4qB locus, the most important finding in our study was detection of DNA methylation rates across entire contracted and normal expanded D4Z4 repeat sequences from the 4qA and 10q loci. As shown in Figure 3, 4qA-derived contracted reads were uniformly hypomethylated in patients with FSHD1, while both 4qA- and 10q-derived reads were uniformly hypomethylated in FSHD2, with the exception of a few reads. These results are similar to those generated in previous studies by Southern blot and bisulfite sequencing analyses (de Greef et al., 2009; Jones et al., 2014; Van Overveld et al., 2003), but our approach allows assessment of focal methylation rate at the nucleotide level. We further analyzed 10q-derived reads in FSHD1, and found that the methylation level was lower at proximal D4Z4 RUs (position 8–13), while it gradually increased (up to _≥_ 60%) at distal RUs (positions 1–7). Given the mimicry of normal expanded 4qA-D4Z4 repeats by 10q-derived reads, these results suggest that only DNA hypermethylation at distal D4Z4 RUs contributes to suppression of the *DUX4* gene in the normal 4qA allele, while contraction of D4Z4 repeats causes hypomethylation of distal D4Z4 similar to proximal D4Z4 in the 10q locus, leading to DUX4 expression and consequent development of FSHD1. Indeed, mean CpG methylation rate of the most distal RUs and disease onset in patients was well-correlated. A larger study of the relationships among methylation rate, D4Z4 contraction, and clinical phenotypes is needed. To this end, we aim to overcome the limitation of decreased acquisition of sequencing reads from alleles with more than 10 RUs.

In this study, we successfully determined the hypomethylation of D4Z4 RUs in individual 4qA fragments in FSHD. The hypomethylation in the contracted D4Z4 in FSHD1 provides a good explanation why the shortening of D4Z4 repeats is associated with severe phenotypes in patients and it induces abnormal DUX4 expression which leads to developing FSHD. Further additional analyses of a large number of patients might give a clue for complete understanding of the pathomechanism of FSHD.

## Data Availability

All data produced in the present study and requests for resources and reagents should be directed to and will be fulfilled by the lead contact, Satoru Noguchi

## Acknowledgments

The authors would like to thank the patients, their families, and physicians who participated in the study.

This study was partly supported by an Intramural Research Grant for Neurological and Psychiatric Disorders of NCNP, under Grant Numbers 3-9 (SN), 2-6 (SN), and 2-5 (IN, SH); AMED, under Grant Number, 22ek0109490h0003 (SN, SH, IN); Nippon Shinyaku Research Grant (YH), KAKENHI (21K15689) (YH), and the FSHD Society (YH).

## Author contributions

Conceptualization, SN; Formal analysis, YH, KI, YS, and SN; Investigation, YH, YK, YS, and MO; Methodology, YH and SN; Patient evaluations, collecting patient samples, and/or clinical data, YS, MM, YO, YT, DK, NA, CM, TM, TH, KN, and KI; Visualization, YH, YS, and SN; Software, KI; Resources, YG and IN; Supervision, SH,

SN, and IN; Project administration, SN; Funding acquisition, YH, SH, SN, and IN; Writing – original draft, YH and SN; Writing – reviewed, all authors.

## Declaration of interests

The authors declare no competing interests.

## STAR Methods

### Human subjects

Samples and data were collected between January 1978 and December 2021 from the National Center of Neurology and Psychiatry registry. Fourteen patients were selected, of whom five had 1, 2, 3, 4, or 5 D4Z4 RUs, while data were inconsistent for seven patients, and two patients showed no bands on linear Southern blotting of genomic DNA samples extracted from peripheral blood lymphocytes. The oldest clinical description available for each patient (data at hospital inspection) was reviewed. Clinical characteristics and the results of Southern blotting are described in Tables 1 and 2, respectively. Materials used in this study were obtained for diagnostic purposes with written informed consent. Fibroblasts from Patient 1 were obtained from the NCNP Biobank. This study was approved by the ethics committee of the National Center of Neurology and Psychiatry, Japan. All participants were enrolled after providing informed consent.

### Genomic DNA preparation

Peripheral blood lymphocytes (10 ml) were combined with 30 ml EL buffer (155 mM NH_4_Cl, 10 mM KHCO_3_, 1 mM EDTA, pH 7.4) on ice for 15 min, followed by centrifugation (KUBOTA 5930, RS-3012M) (840 x g, 10 min, room temperature). After a repeat EL buffer wash, pellets were suspended in 3 ml NL buffer (10 mM Tris-HCl, 2 mM EDTA, 400 mM NaCl, pH 8.2), followed by addition of 1% SDS and proteinase K and incubation at 37°C overnight. DNA lysis solution was added with 1 ml 5 M NaCl, followed by phenol/chloroform extraction and ethanol precipitation. DNA pellets were suspended in TE buffer.

Fibroblasts grown in culture dishes were lysed in 10 mM Tris-HCl, 10 mM EDTA, 150 mM NaCl, pH 8.0 containing 0.5% SDS and proteinase K at 55°C overnight, followed by phenol/chloroform extraction and ethanol precipitation. DNA pellets were suspended in TE buffer.

### DNA library preparation

DNA libraries were prepared using a ligation sequencing kit (Oxford Nanopore Technologies, SQK-LSK109). To generate Cas9 ribonucleoprotein complexes (RNPs), annealed 1 μM tracrRNA-crRNA pool (CR1/CR2/CR3/CR4) and 0.5 μM HiFi Cas9 were incubated at room temperature for 30 min. Genomic DNA (2 μg) was dephosphorylated with Quick Calf Intestinal Phosphatase (NEB, #M0525S) at 37°C for 10 min, followed by 80°C for 2 min. For Cas9 RNP cleavage and dA-tailing, dephosphorylated genomic DNA samples were treated with Cas9 RNPs, Taq polymerase (NEB, #M0273S), and dATP (NEB, #N0440S) at 37°C for 30 min, followed by 72°C for 5 min. For native barcode ligation, native barcoding expansion 1–12 (Oxford Nanopore Technologies, EXP-NBD104) were ligated to cleaved and dA-tailed genomic DNAs using Blunt/TA Ligase Master Mix (NEB, #M0367L) at room temperature for 10 min, followed by purification with Agencourt AMPure XP Beads (Beckman Coulter, #A63880) on a magnet. AMII adapters were ligated to barcoded genomic DNA using Quick T4 DNA ligase (NEB, #E7185A) at room temperature for 10 min, followed by purification with AMPure XP Beads on a magnet. The DNA library from Cas9-targeted native barcoding was primed into a MinION Flow Cell (FLO-MIN106D) on a MinION Mk1C and sequencing was performed for 20–21 h.

The crRNA design tool, CHOPCHOP (Labun et al., 2019), was used to design crRNAs, which were synthesized by Integrated DNA Technologies as follows: CR1, 5’gataccgacagcaatagtcc3’; CR2, 5’gtccttcagcactccacatc3’; CR3, 5’ctataggatccacagggagg3’; and CR4, 5’tgtcaaggtttggcttatag3’.

### Data analysis

Bases were called from Fast5 files using Guppy to generate Fastq files. Alignment to the reference sequence, which contains 10 D4Z4 RUs and flanking sequences from 3950 bp upstream of CR1 to 251 bp downstream of CR4 (Supplemental file 1), was conducted using Minimap2. Reference sequences were constructed using SnapGene software (from Insightful Science; available at snapgene.com). For DNA methylation analysis, sense- and antisense-strand reads from the 4qA and 10q loci were re-aligned to the corresponding reference sequences and then Nanopolish was performed (Simpson et al., 2017). Reference sequences contained the detected size of D4Z4 RUs and flanking sequences from 327 bp downstream of CR2 to 1 bp upstream of CR3. Sequence containing 1 D4Z4 RU is presented as Supplemental file 2. Unipro UGENE free software and Integrative genomics viewer were used for sequence alignment (Okonechnikov et al., 2012; Robinson et al., 2011). For analysis of correlation between the distal D4Z4 CpG methylation rate and clinical symptoms, we calculated mean CpG methylation rates of the most distal D4Z4 RUs (RU3, RU2, and the promoter region of RU1) for all 4qA-reads obtained from each FSHD1 sample. Mean methylation rate or D4Z4 length, and age at disease onset or age at hospital inspection were analyzed and plotted with Graphpad Prism, and correlation coefficients were calculated by linear regression.

## Resource availability

### Lead contact

Further information and requests for resources and reagents should be directed to and will be fulfilled by the lead contact, Satoru Noguchi

### Materials availability

This study did not generate new unique reagents.

### Data and code availability

This study did not generate codes. Any additional information required to reanalyze the data reported in this paper is available from the lead contact upon request.

## Supplementary information

Supplemental information can be found online at

Supplemental Figure 1. Characteristic sequences detected by nCATS. Sequences of representative (A) 4qA- and (B) 10q-derived reads obtained from the indicated samples. The XapI/non-XapI and BlnI/non-BlnI sites in the most distal D4Z4 RU are shown. In Samples 8, 14, and 15, the XapI, XapI and non-BlnI, and non-XapI sites, respectively, in the second most distal D4Z4 RU are shown, due to the difficulty in identifying restriction sites.

Supplemental Table 1. Lengths of reads derived from the 4qA locus in each patient.

Supplemental Table 2. Lengths of reads derived from the 10q locus in each patient.

Supplemental Table 3. Methylation rates across all D4Z4 RUs at the 4qA and 10q loci.

Supplemental file 1. Reference sequence for identification of D4Z4 RUs.

Supplemental file 2. Reference sequence of 1 RU for methylation analysis (sense read).

## Notes

### Competing Interest Statement

The authors have declared no competing interest.

### Author Declarations

This study was approved by the ethics committee of the National Center of Neurology and Psychiatry, Japan. All participants were enrolled after providing informed consent.

