## Supplemental materials for "Hypomethylation of contracted D4Z4 repeats in facioscapulohumeral muscular dystrophy"

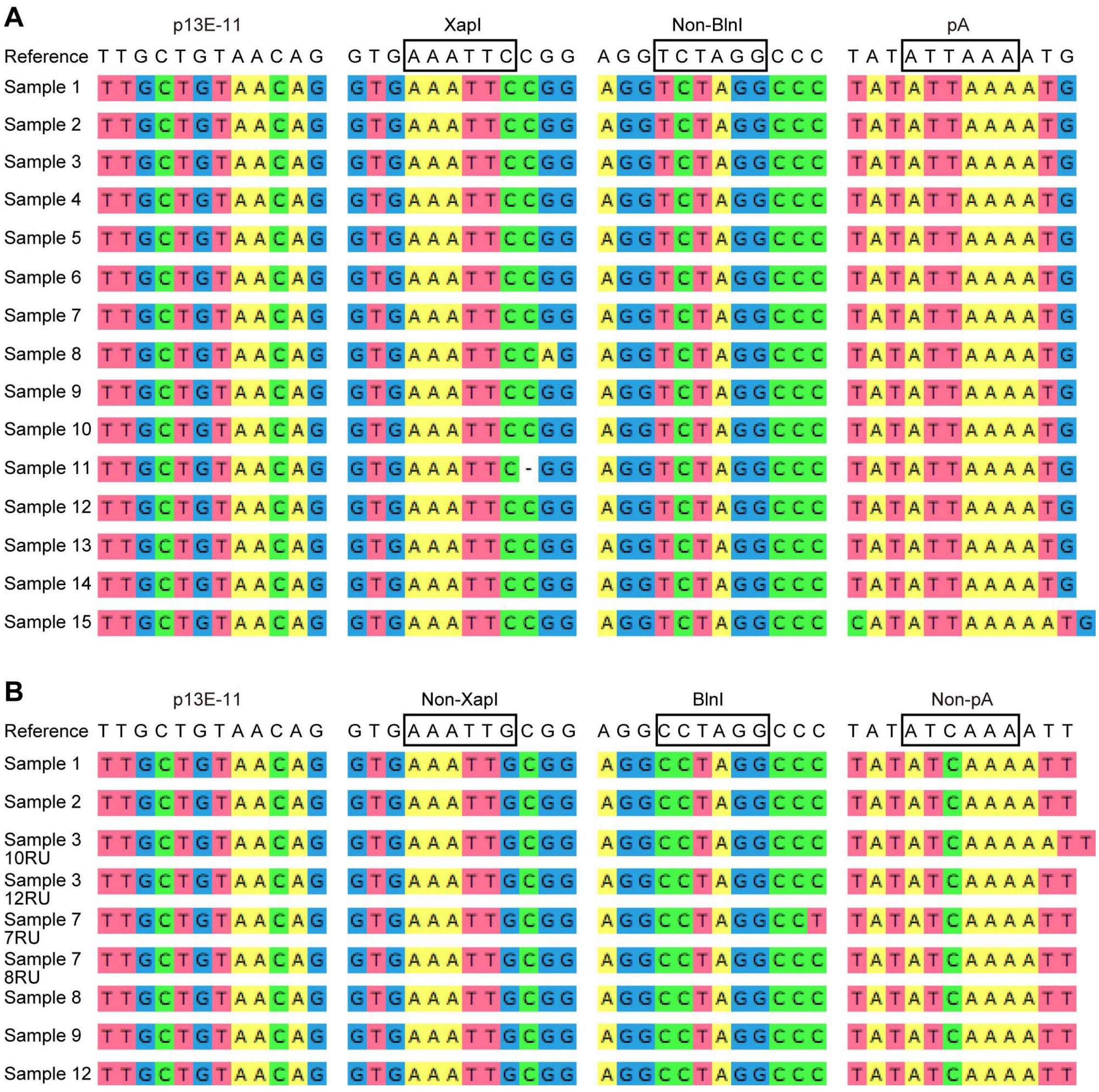


Supplemental Figure 1. Characteristic sequences detected on nCATS. (A) The sequences in the 4qA-derived representative read, and (B) the sequences in the 10q-derived representative read obtained from indicated samples. The XapI/non-XapI and BlnI/non-BlnI sites in the most distal D4Z4 RU are shown. In Samples 8, 14 and 15, the XapI site, the XapI and non-BlnI sites, and the non-XapI site in the second most distal D4Z4 RU, respectively are shown due to the difficulty to identify the restriction sites.

Supplemental Table 1. The length of reads derived from 4qA locus in each patient.

| Sample ID | Direction | Length (bp) |
|---|---|---|
| Sample 1 | AntiSense | 5221 |
| | Sense | 5316 |
| | AntiSense | 5372 |
| | AntiSense | 5390 |
| | Sense | 5391 |
| | Sense | 5397 |
| | Sense | 5425 |
| | AntiSense | 5428 |
| | AntiSense | 5434 |
| | Sense | 5440 |
| | AntiSense | 5449 |
| | Sense | 5469 |
| | Sense | 5794 |
| Sample 2 | Sense | 8535 |
| | AntiSense | 8575 |
| | AntiSense | 8591 |
| | AntiSense | 8611 |
| | Sense | 8619 |
| | AntiSense | 8658 |
| | AntiSense | 8676 |
| | AntiSense | 8679 |
| | Sense | 8681 |
| | AntiSense | 8700 |
| | Sense | 8703 |
| | Sense | 8720 |
| | AntiSense | 8721 |
| | Sense | 8732 |
| | AntiSense | 8976 |
| Sample 3 | AntiSense | 11659 |
| | AntiSense | 11861 |
| | AntiSense | 11872 |
| | AntiSense | 11912 |
| | AntiSense | 11922 |
| | AntiSense | 11923 |
| | AntiSense | 11924 |
| | Sense | 11952 |
| | Sense | 11967 |
| | Sense | 12010 |
| | AntiSense | 11909 |
| Sample 4 | AntiSense | 14566 |
| | Sense | 14863 |
| | Sense | 15102 |
| | AntiSense | 15102 |
| | AntiSense | 15134 |
| | Sense | 15163 |
| | AntiSense | 15165 |
| | AntiSense | 15177 |

| | | |
|---|---|---|
| | AntiSense | 15185 |
| | AntiSense | 15217 |
| | AntiSense | 15466 |
| Sample 5 | Sense | 18383 |
| | AntiSense | 18397 |
| | AntiSense | 18404 |
| | AntiSense | 18448 |
| | Sense | 18453 |
| | Sense | 18730 |
| Sample 6 | AntiSense | 5281 |
| | Sense | 5337 |
| | AntiSense | 5735 |
| | Sense | 5737 |
| | Sense | 5742 |
| | AntiSense | 5756 |
| Sample 7 | AntiSense | 37844 |
| | AntiSense | 37903 |
| | Sense | 37923 |
| | AntiSense | 37926 |
| | Sense | 37959 |
| Sample 8 | Sense | 44517 |
| Sample 9 | AntiSense | 14511 |
| | AntiSense | 14792 |
| | AntiSense | 14851 |
| | AntiSense | 14903 |
| | Sense | 14951 |
| | Sense | 15044 |
| | Sense | 15133 |
| | Sense | 15157 |
| Sample 10 | Sense | 18042 |
| | Sense | 18067 |
| | AntiSense | 18079 |
| | AntiSense | 18151 |
| | Sense | 18313 |
| | AntiSense | 18396 |
| | Sense | 18425 |
| | AntiSense | 18434 |
| | AntiSense | 18452 |
| | AntiSense | 18466 |
| | AntiSense | 18694 |
| | Sense | 18756 |
| Sample 11 | Sense | 18450 |
| | Sense | 18460 |
| | Sense | 18412 |
| Sample 12 | Sense | 18480 |
| | AntiSense | 18513 |
| | AntiSense | 18607 |
| | Sense | 18355 |
| | Sense | 12020 |

| | | |
|---|---|---|
| Sample 13 | Sense | 11873 |
| | Sense | 11838 |
| | AntiSense | 11838 |
| | AntiSense | 10505 |
| Sample 14 | AntiSense | 10520 |
| Sample 15 | Sense | 11596 |
| | AntiSense | 11878 |

Supplemental Table 2. The length of reads derived from 10q locus in each patient.

| Sample ID | Direction | Length (bp) |
|---|---|---|
| Sample 1 | Sense | 44383 |
| | Antisense | 44837 |
| Sample 2 | Antisense | 43595 |
| | Antisense | 43780 |
| | Sense | 44612 |
| Sample 3 | Sense | 34791 |
| | Sense | 34825 |
| | Sense | 41183 |
| | Sense | 41390 |
| Sample 7 | Sense | 24845 |
| | Sense | 24872 |
| | Antisense | 24905 |
| | Sense | 24980 |
| | Antisense | 25002 |
| | Sense | 28081 |
| | Antisense | 28116 |
| | Sense | 28194 |
| | Antisense | 28246 |
| | Sense | 28301 |
| | Antisense | 28335 |
| Sample 8 | Sense | 41213 |
| Sample 9 | Antisense | 37844 |
| | Antisense | 37871 |
| | Antisense | 37879 |
| | Sense | 38166 |
| | Sense | 38487 |
| Sample 12 | Sense | 41111 |
| | Sense | 41368 |

Supplemental Table 3. The methylation rate at the

| Sample ID | locus | Methylation rate (%) | | | | | | | | | | | | | |
|---|---|---|---|---|---|---|---|---|---|---|---|---|---|---|---|
| | | D4Z4 | | | | | | | | | | | | | |
| | | 13 | 12 | 11 | 10 | 9 | 8 | 7 | 6 | 5 | 4 | 3 | 2 | 1 | |
| | | | | | | | | | | | | | | Promoter | Gene body |
| Sample 1 | 4qA | | | | | | | | | | | | | 0.9 | 4.4 |
| | | | | | | | | | | | | | | 1.5 | 4.3 |
| | | | | | | | | | | | | | | 1.6 | 0.0 |
| | | | | | | | | | | | | | | 0.0 | 3.4 |
| | | | | | | | | | | | | | | 4.4 | 0.0 |
| | | | | | | | | | | | | | | 0.7 | 1.1 |
| | | | | | | | | | | | | | | 0.0 | 3.7 |
| | | | | | | | | | | | | | | 0.8 | 2.7 |
| | | | | | | | | | | | | | | 14.1 | 30.5 |
| | | | | | | | | | | | | | | 1.7 | 1.0 |
| | | | | | | | | | | | | | | 4.2 | 6.8 |
| | | | | | | | | | | | | | | 4.7 | 1.3 |
| | | | | | | | | | | | | | | 7.1 | 4.2 |
| | 10q | 22.7 | 8.8 | 10.1 | 24.4 | 13.8 | 18.9 | 40.6 | 35.3 | 48.4 | 44.9 | 66.0 | 66.1 | 56.5 | 36.4 |
| | | 4.1 | 15.7 | 14.3 | 12.7 | 51.7 | 24.5 | 13.2 | 24.3 | 27.0 | 56.4 | 53.8 | 56.1 | 57.6 | 57.5 |
| Sample 2 | 4qA | | | | | | | | | | | | 8.5 | 8.0 | 16.9 |
| | | | | | | | | | | | | | 6.4 | 7.1 | 14.7 |
| | | | | | | | | | | | | | 9.1 | 30.9 | 55.0 |
| | | | | | | | | | | | | | 1.7 | 10.5 | 11.9 |
| | | | | | | | | | | | | | 2.8 | 3.4 | 1.0 |
| | | | | | | | | | | | | | 5.6 | 22.1 | 3.9 |
| | | | | | | | | | | | | | 8.6 | 16.7 | 54.0 |
| | | | | | | | | | | | | | 4.5 | 11.4 | 19.4 |
| | | | | | | | | | | | | | 13.2 | 19.6 | 72.0 |
| | | | | | | | | | | | | | 2.8 | 17.6 | 23.9 |
| | | | | | | | | | | | | | 7.1 | 13.1 | 14.3 |
| | | | | | | | | | | | | | 0.8 | 3.1 | 4.8 |
| | | | | | | | | | | | | | 6.1 | 15.0 | 4.8 |
| | | | | | | | | | | | | | 16.8 | 22.8 | 56.3 |
| | | | | | | | | | | | | | 4.1 | 7.4 | 17.7 |
| | 10q | 13.0 | 18.8 | 21.5 | 22.1 | 18.5 | 17.9 | 34.0 | 26.9 | 39.7 | 36.2 | 55.9 | 64.0 | 43.3 | 82.5 |
| | | 17.5 | 21.8 | 47.1 | 47.6 | 80.3 | 52.8 | 57.3 | 86.5 | 92.5 | 80.6 | 69.8 | 72.5 | 66.7 | 84.8 |
| | | 18.9 | 8.1 | 27.8 | 40.0 | 47.3 | 79.6 | 38.2 | 47.1 | 56.7 | 65.5 | 51.8 | 74.6 | 53.2 | 94.8 |
| Sample 3 | 4qA | | | | | | | | | | | 0.7 | 3.0 | 5.8 | 1.9 |
| | | | | | | | | | | | | 2.8 | 3.7 | 6.1 | 39.1 |
| | | | | | | | | | | | | 1.3 | 2.7 | 9.4 | 2.4 |
| | | | | | | | | | | | | 5.6 | 0.4 | 2.8 | 0.0 |
| | | | | | | | | | | | | 3.7 | 6.9 | 3.2 | 25.9 |
| | | | | | | | | | | | | 3.5 | 1.2 | 1.7 | 29.4 |
| | | | | | | | | | | | | 3.3 | 7.9 | 4.3 | 4.8 |
| | | | | | | | | | | | | 1.3 | 4.4 | 0.0 | 0.0 |
| | | | | | | | | | | | | 8.8 | 8.9 | 11.8 | 12.9 |
| | | | | | | | | | | | | 0.6 | 1.3 | 2.4 | 8.1 |
| | | | | | | | | | | | | 1.4 | 2.5 | 1.9 | 11.3 |
| | 10q | | | | 8.2 | 10.0 | 14.8 | 16.0 | 11.6 | 14.7 | 21.6 | 20.0 | 34.8 | 18.3 | 38.7 |
| | | | | | 45.5 | 66.0 | 63.4 | 74.6 | 81.0 | 87.0 | 89.3 | 93.1 | 92.9 | 77.9 | 92.9 |
| | | | 15.5 | 13.2 | 15.6 | 9.5 | 11.9 | 19.1 | 36.6 | 18.7 | 15.0 | 18.6 | 47.4 | 26.1 | 40.0 |
| | | | 11.3 | 6.8 | 5.5 | 6.1 | 17.2 | 16.3 | 10.7 | 12.5 | 11.2 | 9.1 | 16.8 | 31.6 | 50.0 |
| | | | | | | | | | | | 0.0 | 4.8 | 8.2 | 13.3 | 40.5 |
| | | | | | | | | | | | 6.1 | 18.5 | 16.9 | 26.5 | 24.5 |
| | | | | | | | | | | | 3.5 | 1.9 | 3.7 | 11.4 | 30.6 |
| | | | | | | | | | | | 0.4 | 7.8 | 1.6 | 0.8 | 4.1 |

| | | | | | | | | | | | | | | | |
|---|---|---|---|---|---|---|---|---|---|---|---|---|---|---|---|
| Sample 4 | 4qA | | | | | | | | | | 2.8 | 2.6 | 4.7 | 5.6 | 2.7 |
| | | | | | | | | | | | 1.4 | 2.2 | 4.2 | 5.9 | 18.8 |
| | | | | | | | | | | | 4.7 | 4.0 | 3.7 | 2.8 | 20.5 |
| | | | | | | | | | | | 3.5 | 9.4 | 2.6 | 4.3 | 17.9 |
| | | | | | | | | | | | 4.1 | 4.8 | 11.7 | 4.3 | 8.7 |
| | | | | | | | | | | | 0.4 | 1.7 | 2.3 | 6.7 | 17.6 |
| | | | | | | | | | | | 3.8 | 2.0 | 3.9 | 1.9 | 1.3 |
| Sample 5 | 4qA | | | | | | | | | 3.1 | 1.7 | 4.5 | 2.8 | 0.9 | 8.7 |
| | | | | | | | | | | 3.1 | 4.1 | 8.0 | 6.7 | 10.3 | 27.5 |
| | | | | | | | | | | 2.9 | 15.9 | 9.9 | 4.0 | 8.0 | 11.2 |
| | | | | | | | | | | 9.1 | 13.7 | 4.4 | 6.5 | 3.9 | 36.9 |
| | | | | | | | | | | 13.7 | 17.0 | 5.6 | 10.7 | 12.6 | 58.4 |
| | | | | | | | | | | 12.2 | 25.5 | 10.4 | 6.8 | 19.2 | 60.0 |
| Sample 6 | 4qA | | | | | | | | | | | | | 0.0 | 0.0 |
| | | | | | | | | | | | | | | 1.7 | 0.0 |
| | | | | | | | | | | | | | | 0.8 | 2.0 |
| | | | | | | | | | | | | | | 0.9 | 0.0 |
| | | | | | | | | | | | | | | 1.7 | 6.0 |
| | | | | | | | | | | | | | | 3.2 | 1.0 |
| Sample 7 | 4qA | | | 6.6 | 4.4 | 2.4 | 6.9 | 6.1 | 8.6 | 6.2 | 13.7 | 4.4 | 17.1 | 21.8 | 60.9 |
| | | | | 4.2 | 6.7 | 2.2 | 0.9 | 4.8 | 2.8 | 1.8 | 0.5 | 2.8 | 11.2 | 10.6 | 5.6 |
| | | | | 4.1 | 5.0 | 2.3 | 7.0 | 3.8 | 3.5 | 4.5 | 6.4 | 15.2 | 12.2 | 7.4 | 10.0 |
| | | | | 1.2 | 4.0 | 3.6 | 1.3 | 5.1 | 10.3 | 9.2 | 3.1 | 3.9 | 8.9 | 0.9 | 5.4 |
| | | | | 1.7 | 5.1 | 1.3 | 6.4 | 1.9 | 6.3 | 6.0 | 2.6 | 8.9 | 11.6 | 15.4 | 45.8 |
| | 10q | | | | | | | 2.8 | 1.4 | 2.1 | 4.8 | 3.0 | 6.8 | 2.8 | 12.8 |
| | | | | | | | | 5.2 | 3.1 | 2.4 | 2.4 | 4.9 | 3.9 | 2.4 | 5.4 |
| | | | | | | | | 3.8 | 2.6 | 3.3 | 3.7 | 1.3 | 8.4 | 6.7 | 1.5 |
| | | | | | | | | 3.6 | 6.8 | 7.0 | 6.2 | 8.0 | 4.8 | 8.7 | 14.3 |
| | | | | | | | | 1.5 | 5.1 | 3.4 | 4.9 | 4.6 | 5.3 | 7.5 | 51.3 |
| | | | | | | | 17.9 | 15.9 | 6.4 | 15.5 | 33.1 | 24.4 | 42.9 | 47.8 | 72.7 |
| | | | | | | | 3.4 | 2.0 | 2.3 | 11.6 | 12.2 | 6.9 | 5.3 | 8.0 | 16.4 |
| | | | | | | | 7.3 | 4.8 | 6.3 | 11.7 | 16.3 | 5.5 | 50.0 | 37.5 | 24.1 |
| | | | | | | | 2.8 | 3.5 | 2.9 | 7.2 | 9.0 | 6.0 | 7.2 | 7.6 | 16.7 |
| | | | | | | | 7.2 | 2.5 | 2.8 | 8.4 | 2.4 | 5.4 | 9.6 | 10.1 | 17.9 |
| | | | | | | | 9.0 | 14.3 | 15.4 | 12.4 | 11.6 | 18.2 | 8.5 | 31.3 | 42.4 |
| Sample 8 | 4qA | 3.6 | 3.8 | 3.7 | 4.2 | 3.6 | 9.1 | 3.5 | 12.4 | 8.2 | 4.8 | 5.6 | 11.7 | 6.7 | 13.6 |
| | 10q | | 4.3 | 7.9 | 10.6 | 8.3 | 6.5 | 13.3 | 3.8 | 13.3 | 9.0 | 3.1 | 9.5 | 7.1 | 8.6 |
| Sample 9 | 4qA | | | | | | | | | | 8.8 | 6.1 | 10.6 | 30.4 | 8.3 |
| | | | | | | | | | | | 2.3 | 0.8 | 0.6 | 3.7 | 0.0 |
| | | | | | | | | | | | 1.2 | 1.3 | 2.0 | 15.2 | 51.7 |
| | | | | | | | | | | | 2.6 | 2.5 | 2.2 | 2.6 | 7.5 |
| | | | | | | | | | | | 0.0 | 4.0 | 8.5 | 8.6 | 37.7 |
| | | | | | | | | | | | 6.5 | 5.4 | 7.5 | 5.7 | 15.7 |
| | | | | | | | | | | | 0.5 | 4.8 | 0.0 | 4.7 | 0.0 |
| | | | | | | | | | | | 2.1 | 3.3 | 2.2 | 2.3 | 1.6 |
| | 10q | | | 12.1 | 7.1 | 14.2 | 15.4 | 30.6 | 15.7 | 22.3 | 35.2 | 22.8 | 33.1 | 10.1 | 40.0 |
| | | | | 24.8 | 29.5 | 35.9 | 43.6 | 20.1 | 20.9 | 22.8 | 41.8 | 47.7 | 41.1 | 71.0 | 84.6 |
| | | | | 11.6 | 13.0 | 21.1 | 45.6 | 35.7 | 30.4 | 29.8 | 44.3 | 53.5 | 59.8 | 60.0 | 72.9 |
| | | | | 12.8 | 16.2 | 17.7 | 12.6 | 16.1 | 20.0 | 20.9 | 40.0 | 33.3 | 46.0 | 19.7 | 52.8 |
| | | | | 13.9 | 19.3 | 29.1 | 25.5 | 35.0 | 16.1 | 14.1 | 31.5 | 28.2 | 36.8 | 14.0 | 65.8 |
| Sample 10 | 4qA | | | | | | | | | 0.6 | 0.9 | 1.5 | 7.2 | 8.3 | 14.6 |
| | | | | | | | | | | 3.7 | 7.1 | 12.4 | 37.6 | 21.4 | 38.5 |
| | | | | | | | | | | 3.9 | 5.6 | 1.5 | 22.2 | 18.9 | 24.7 |
| | | | | | | | | | | 4.0 | 2.5 | 6.5 | 9.9 | 17.4 | 26.7 |
| | | | | | | | | | | 2.6 | 10.0 | 2.7 | 5.1 | 11.1 | 9.9 |
| | | | | | | | | | | 4.0 | 1.8 | 2.7 | 9.3 | 16.5 | 32.1 |
| | | | | | | | | | | 1.6 | 0.9 | 3.8 | 10.6 | 8.8 | 19.7 |

| | | | | | | | | | | | | | | | |
|---|---|---|---|---|---|---|---|---|---|---|---|---|---|---|---|
| | | | | | | | | | | 0.8 | 2.4 | 10.5 | 7.3 | 31.3 | 3.8 |
| | | | | | | | | | | 2.7 | 6.5 | 8.9 | 8.3 | 23.0 | 44.0 |
| | | | | | | | | | | 2.7 | 8.4 | 4.0 | 6.9 | 12.3 | 17.9 |
| | | | | | | | | | | 6.0 | 11.8 | 11.3 | 27.3 | 15.5 | 71.4 |
| | | | | | | | | | | 4.9 | 8.8 | 12.4 | 17.3 | 10.4 | 42.6 |
| Sample 11 | 4qA | | | | | | | | | 6.4 | 8.9 | 8.2 | 18.8 | 23.1 | 43.4 |
| | | | | | | | | | | 7.0 | 3.5 | 7.0 | 19.7 | 18.7 | 60.3 |
| | | | | | | | | | | 13.6 | 14.7 | 25.9 | 38.6 | 34.8 | 95.9 |
| Sample 12 | 4qA | | | | | | | | | 4.6 | 4.8 | 3.2 | 11.6 | 7.0 | 57.0 |
| | | | | | | | | | | 8.4 | 9.2 | 17.0 | 9.3 | 12.8 | 19.5 |
| | | | | | | | | | | 3.7 | 2.5 | 3.2 | 6.0 | 14.4 | 10.6 |
| | | | | | | | | | | 1.9 | 2.6 | 3.0 | 3.0 | 7.7 | 0.0 |
| | 10qA | | 5.8 | 41.0 | 33.6 | 22.0 | 18.7 | 49.4 | 59.3 | 47.0 | 79.7 | 61.2 | 82.4 | 79.3 | 91.5 |
| | | | 9.5 | 21.5 | 21.3 | 34.0 | 27.5 | 38.1 | 24.8 | 38.1 | 33.1 | 29.5 | 47.2 | 60.0 | 87.5 |
| Sample 13 | 4qA | | | | | | | | | | | 3.5 | 0.8 | 1.6 | 8.9 |
| | | | | | | | | | | | | 1.2 | 7.8 | 2.9 | 12.3 |
| | | | | | | | | | | | | 13.5 | 22.4 | 17.6 | 39.2 |
| | | | | | | | | | | | | 31.0 | 17.3 | 15.8 | 83.6 |
| | | | | | | | | | | | | 1.4 | 1.3 | 4.7 | 1.2 |
| Sample 14 | 4qA | | | | | | | | | | | 15.6 | 16.4 | 25.7 | 21.1 |
| Sample 15 | 4qA | | | | | | | | | | | 1.3 | 16.9 | 4.5 | 15.8 |
| | | | | | | | | | | | | 5.2 | 7.8 | 51.6 | 41.7 |
